# COVID-19 Pandemic in University Hospital: Is There an Effect on The Medical Interns?

**DOI:** 10.1101/2020.10.01.20205112

**Authors:** WeiHonn Lim, Li Ying Teoh, Kanesh Kumaran A/L Seevalingam, Shanggar Kuppusamy

## Abstract

**Introduction:** Coronavirus Disease 2019 (COVID-19) pandemic has disrupted the current healthcare system and carries a major impact to the healthcare workers (HCW). University Malaya Medical Centre (UMMC) has been selected as one of the centres in managing COVID-19 cases in Malaysia. Many HCW including the medical interns, are directly or indirectly involved in the management.

**Methods:** This is a cross-sectional, pilot study to determine the impact of the pandemic on UMMC medical interns. A survey which comprises 37-items was used. Data are analysed by Ordinal Logistic Regression Analysis.

**Results:** Our study shows that medical interns are tired (p = 0.014), starving (p = 0.004), have inadequate exercises (p = 0.004) and burdened with heavy workload (p=0.023) during pandemic period. Many are depressed (p = 0.043), scared to work (p = 0.03), and worried of getting infected (p < 0.05). Some quarrel with their colleagues (p < 0.05), losing contact with friends (p = 0.022) and feel that it will be beneficial to have a peer support group (p = 0.027).

**Conclusion:** In summary, the impact of COVID-19 amongst medical interns is significant and their overall well-being should be protected without jeopardising their training.

## Introduction

Since the outbreak of COVID-19, this novel virus has invaded many countries and exerts a great impact in the current healthcare system.[1, 2] Limited medical resources against an unpredictable disease burden the health system capacity.[3, 4] Furthermore, many HCW have been exhausted from an overwhelming workload globally. Malaysia experiences a similar fate starting from March 2020. In response to this pandemic crisis, Malaysia has implemented Movement Control Order (MCO) from 18th March 2020 in several stages, and currently we are in the Recovery Period of MCO.[5]

The medical interns, also named as the House Officers (HO) in Malaysia, comprise of medical graduates who will require two years of medical training. During this period of training, they rotate to six compulsory clinical postings such as Internal Medicine, Surgery, Obstetrics and Gynaecology, Orthopaedic and Paediatric departments. Addition to that is a choice of a posting in either Psychiatry, Emergency Medicine or Anaesthesiology department. [6] They are required to complete at least 4 months rotation in each department with a certain expectation in their clinical exposure to fulfil, under full supervision by their specialists and consultants. These two years of training are tough and medical interns must withstand a great level of work stress while trying to gain more clinical experiences during the posting in each department.

As a teaching university hospital in Malaysia, UMMC has been gazetted as one of the designated COVID centres. To fulfil its role, many changes have been adapted to accommodate the surge of COVID cases: from rescheduling clinic appointments and elective operations to deployment of more HCW to Emergency Department as “frontliners” and gazetting more designated COVID wards to manage infected cases. [7, 8]

For medical interns to work during pandemic periods, there are few challenges encountered: a sudden rise in workload which strains their physical fitness; fear of contracting COVID-19 and worrying of unable to complete their medical training on-time exerts an immense pressure to their psychology; conflict between colleagues due to sudden change of workload and losing contact with their friends, families and loved ones during this COVID-19 season affects them indirectly. [4, 8-11] Additionally, they are required to complete their tasks with high expectation from their supervisors as they are expected to perform above par in order to become a safe doctor in the community. To protect oneself from this contagious disease, they must heighten their vigilance during their works. All these invisible hurdles not only affect their mental states but their performances. Nevertheless, the degree of impact from this pandemic on the medical interns needs to be addressed to ensure their well-beings are protected.

## Methods

A survey form is created and drafted in English language which composed of two components: demographic status (gender, ethnics, age, number of posting and qualification attained) and the questionnaire itself which comprises of five domains: physical health, emotion/ psychological coping, social/ relationship handling, medical training progression and preparedness in handling a pandemic situation. The questionnaire uses a five-response Likert Scale: ranging from 1 to 5 which represent totally disagree, disagree, neutral, agree and strongly agree respectively.

The questionnaire is uploaded to an online survey form - Google Form and a link is generated. The invitation for participation in the study is sent by email and electronic devices via WhatsApp, with the link to assess the questionnaire. Electronic Participant Information Sheets (PIS) are provided in the first page of the survey form. All participations are self-voluntary whereby they can choose not to participate if they have disagreed to include their responses in this study result. The questionnaire is self-administered by the participants in anonymity. Participation is opened from April 2020 to June 2020.

Subsequently, all data are downloaded into Excel form and transcript into SPSS Version 16.0. Demographic data are summarised by descriptive analysis. Association between demographic factors and each item in the questionnaire is analysed by using Ordinal Logistic Regression Analysis. Subsequently, data are analysed using Generalised Linear Models where demographic data were entered as the factor and all items in the questionnaire were keyed in as dependent variables. For each factor, groups with majority numbers are made as comparators to avoid potential bias: Female, Malay ethnic and local graduates are listed as comparators to others in their respective groups.

## Results

There are total of 236 responses recorded in our Google Form. Out of these numbers, 232 participants consented to join which contributes 98.3% of response rate while only four of them declined to participate. Of 232 participants who agreed to participate in this survey, majority are females (62.1%) and of Malay ethnics (45.3%). Most of them attained their medical degrees from local university (60.8%) and had completed at least 3 postings (56.9%).

**Table 1:** Basic Demographic Data of Medical Interns

| *Variables* | *Male*<br>*(n = 88)* | *Female*<br>*(n = 144)* | *Total*<br>*(n = 232)* |
| --- | --- | --- | --- |
| **Age, years (SD)** | **27 (1.38)** | **27 (1.74)** | **27 (1.6)** |
| **Ethnics, n (%)** |  |  |  |
| Malay | 33 (14.2) | 72 (31.0) | 105 (45.3) |
| Chinese | 35 (15.1) | 43 (18.5) | 78 (33.6) |
| Indian | 17 (7.3) | 24 (10.3) | 41 (17.7) |
| Others | 3 (1.3) | 5 (2.2) | 8 (3.4) |
| **Qualification Attained, n (%)** |  |  |  |
| Local University | 56 (24.1) | 85 (36.6) | 141 (60.8) |
| Overseas University | 18 (7.8) | 44 (19.0) | 62 (26.7) |
| Twinning Programmes | 14 (6.0) | 15 (6.5) | 29 (12.5) |
| **Number of Posting, n (%)** |  |  |  |
| 1 | 13 (5.6) | 14 (6.0) | 27 (11.6) |
| 2 | 14 (6.0) | 25 (10.8) | 39 (16.8) |
| 3 | 24 (10.3) | 42 (18.1) | 66 (28.4) |
| 4 | 20 (8.6) | 36 (15.5) | 56 (24.1) |
| 5 | 6 (2.6) | 11 (4.7) | 17 (7.3) |
| 6 | 11 (4.7) | 16 (6.9) | 27 (11.6) |

**Table 2:**
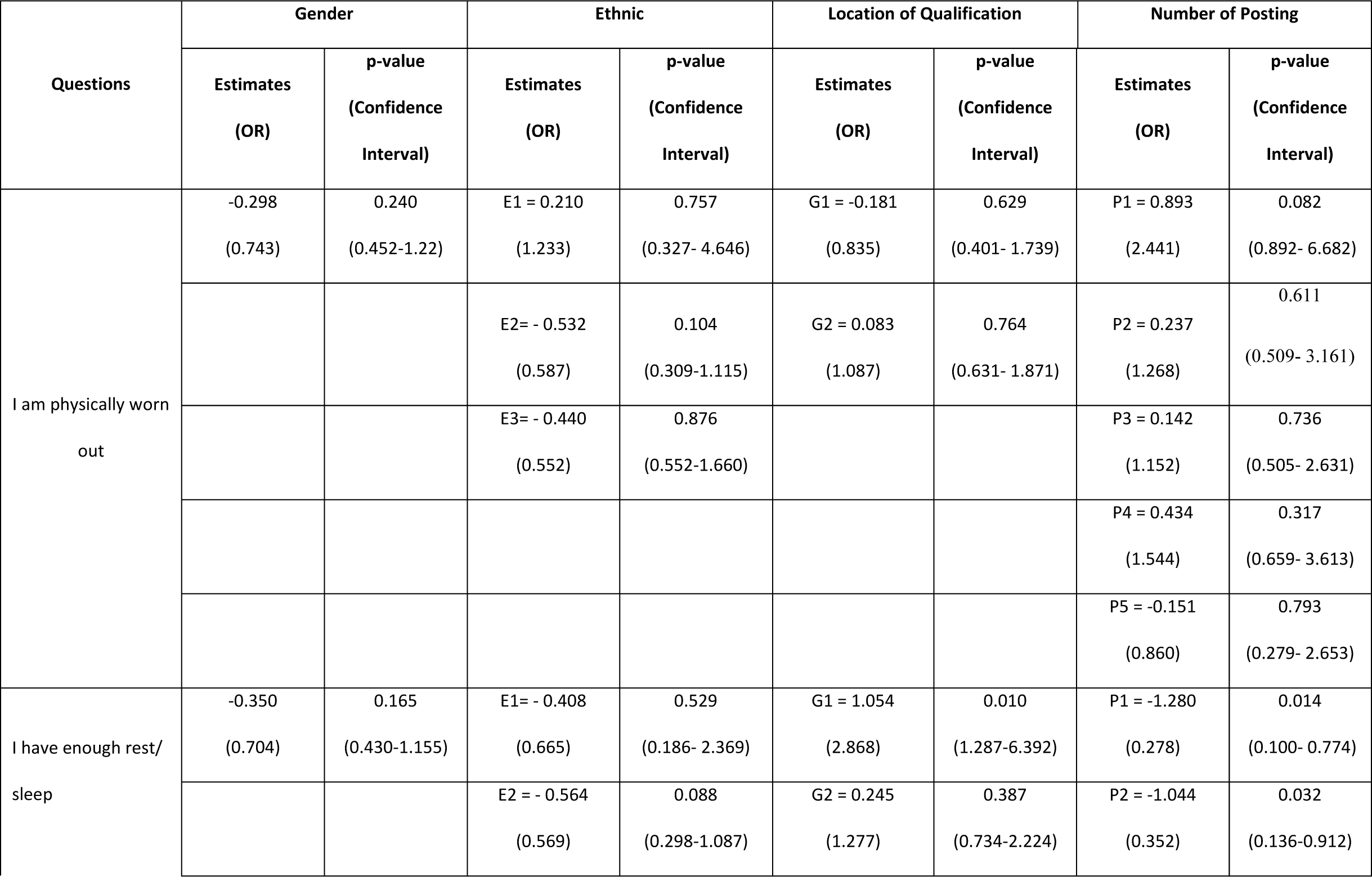
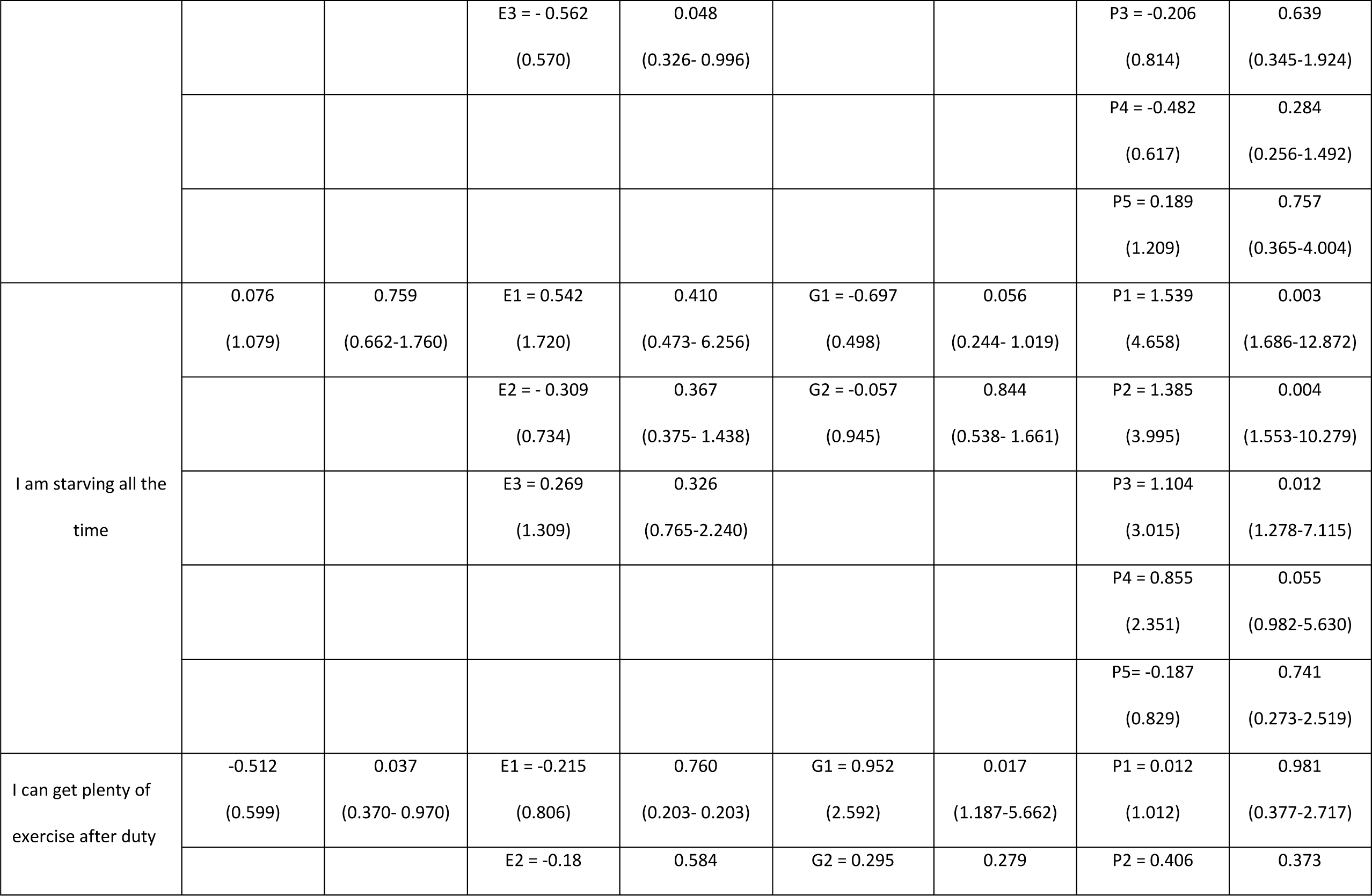
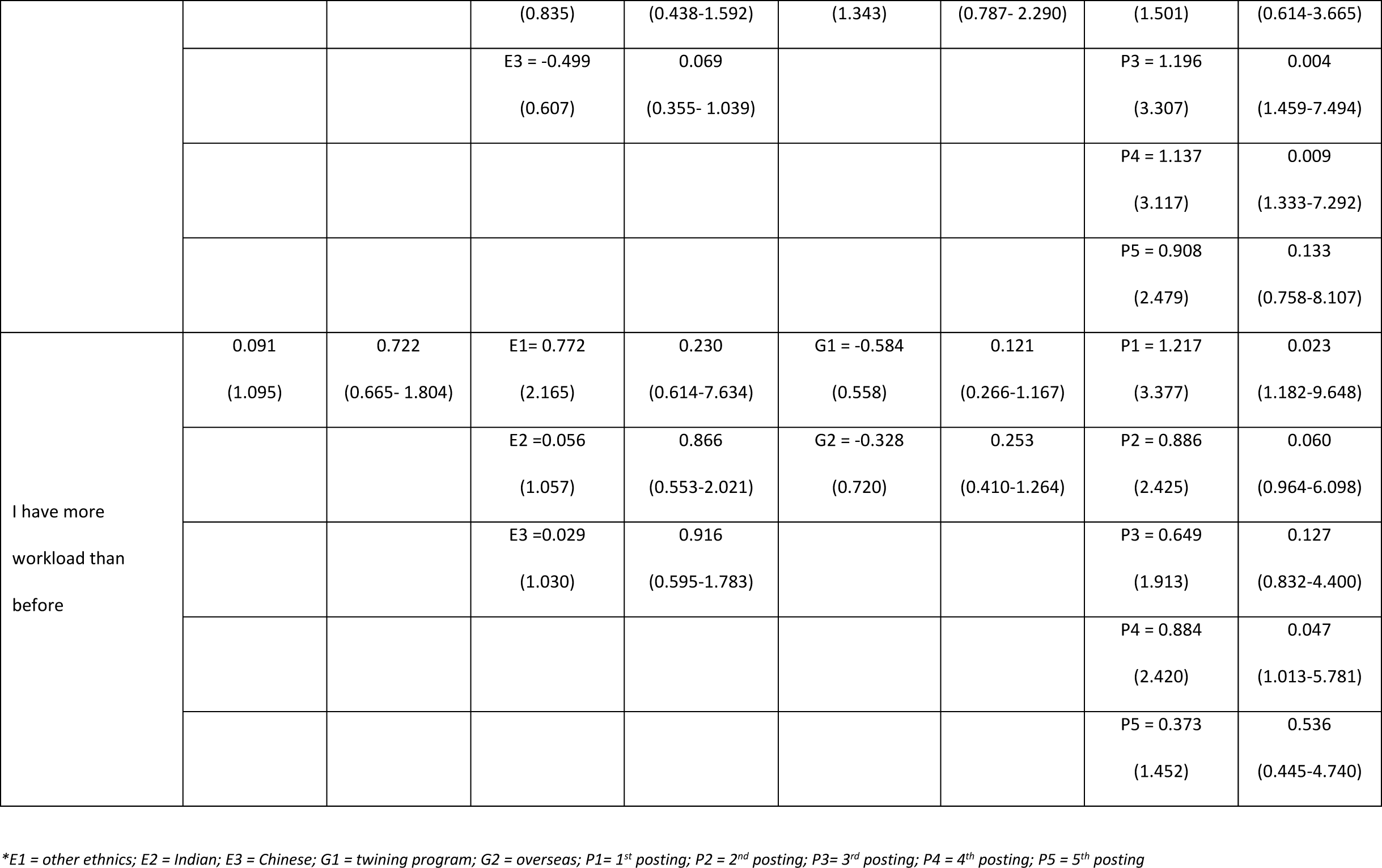
Association between Demographic Factors with Physical Health

**Table 3:**
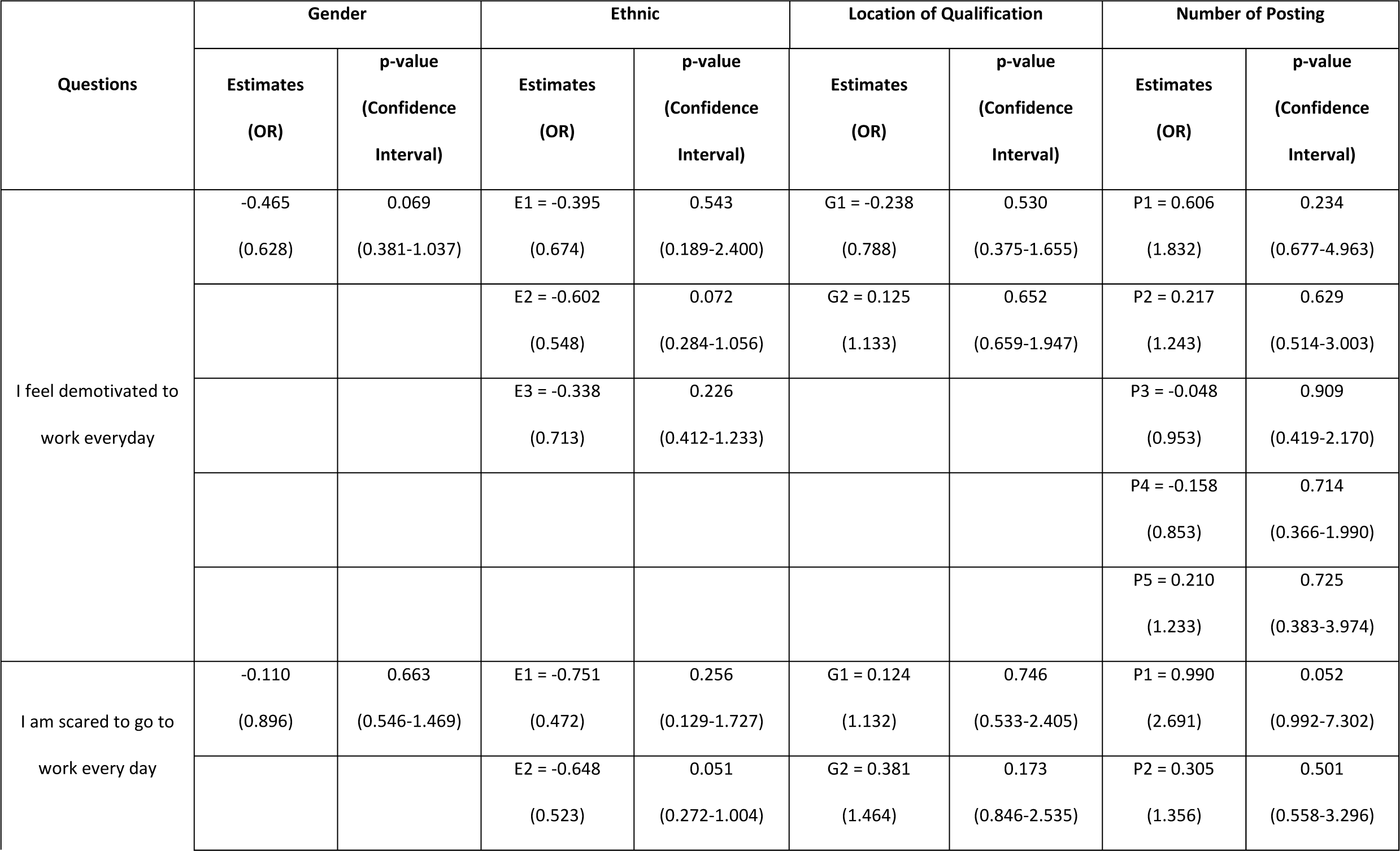
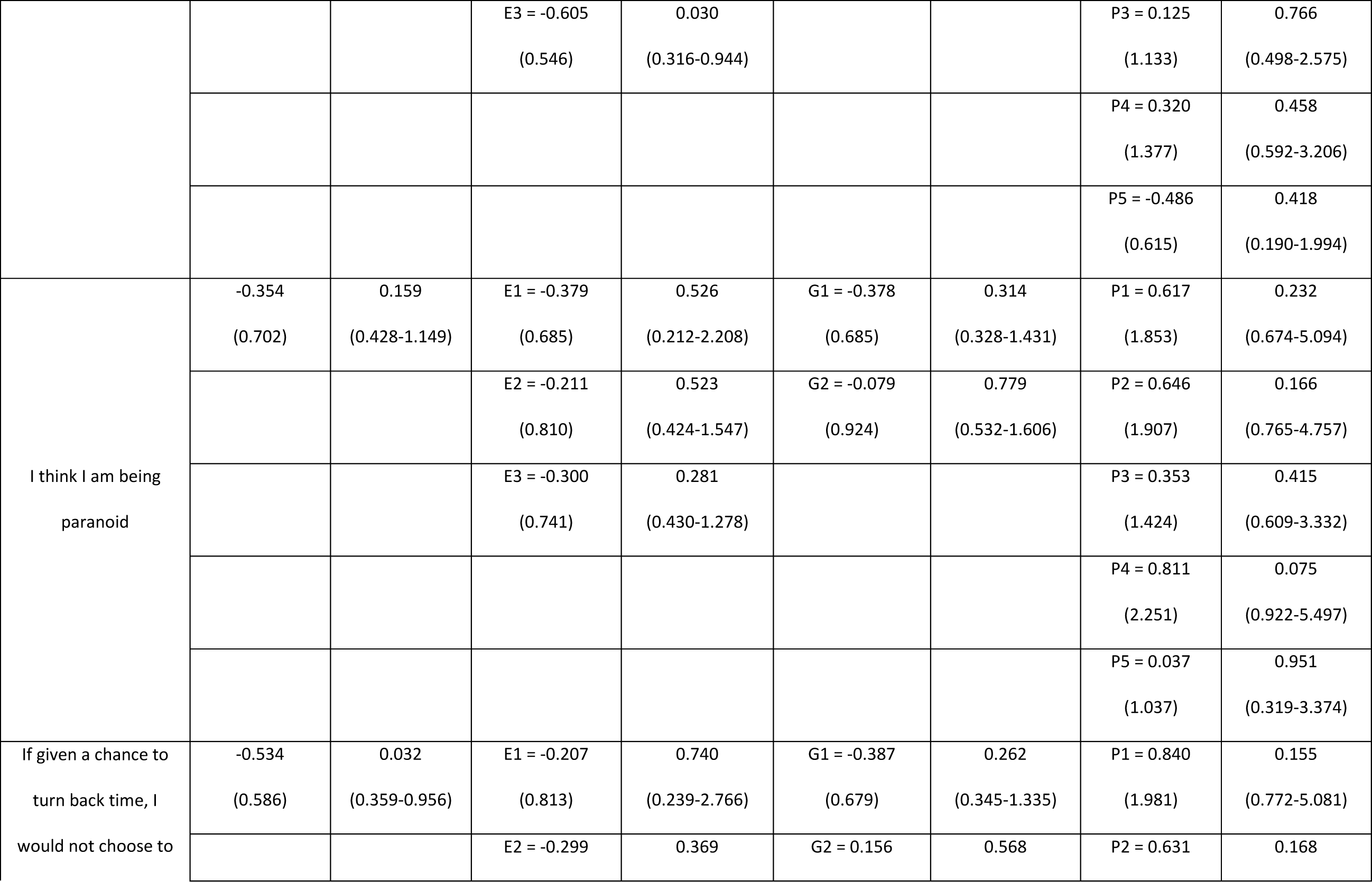
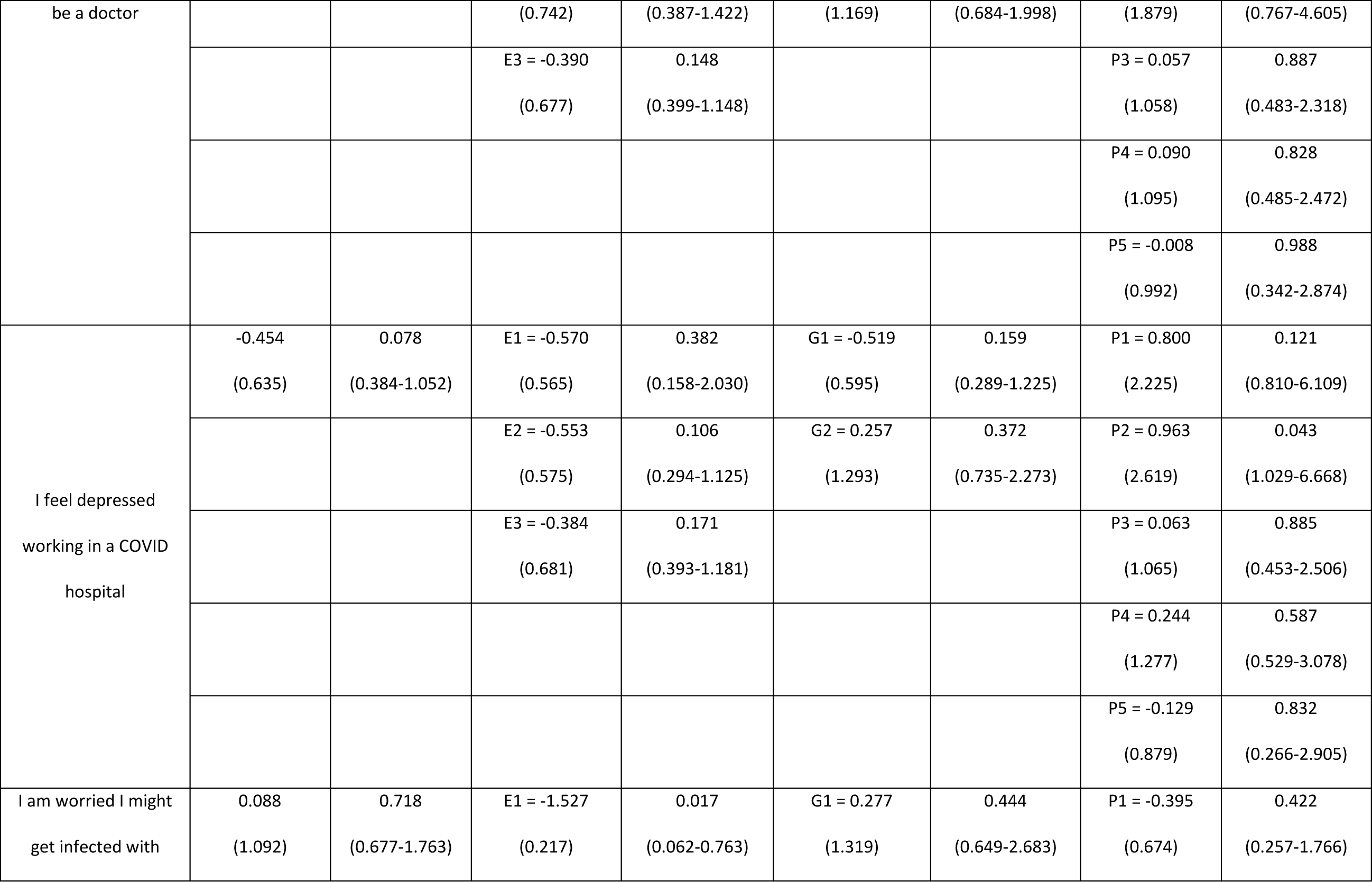
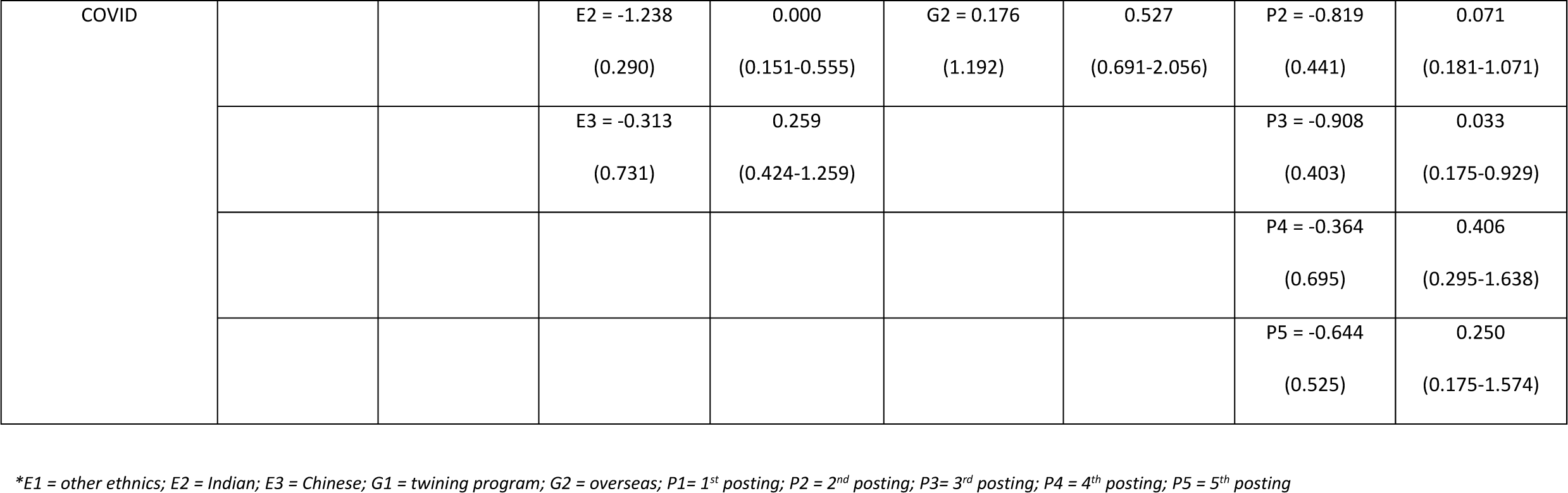
Association between Demographic Factors with Emotion/ Psychological Coping

**Table 4:**
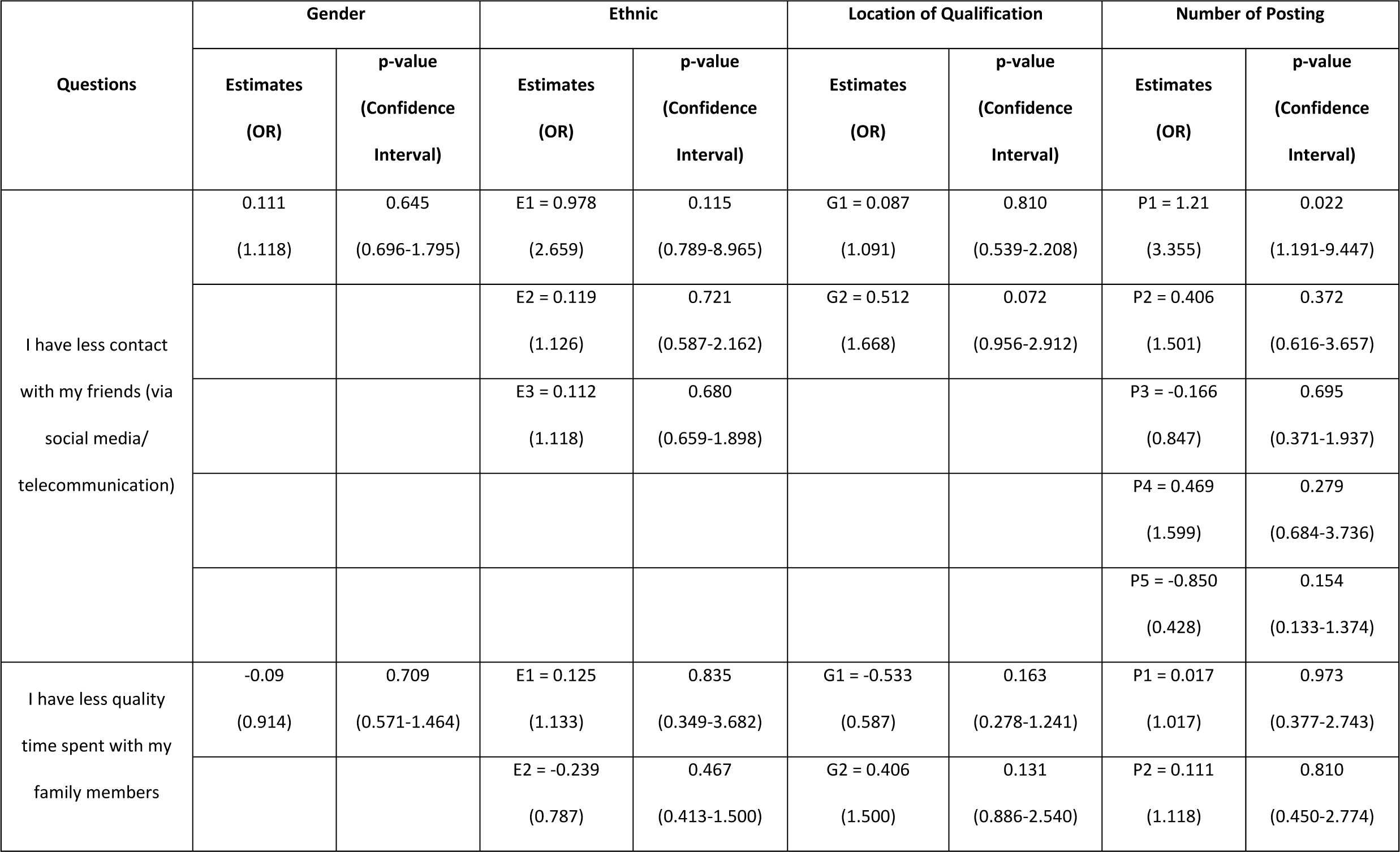
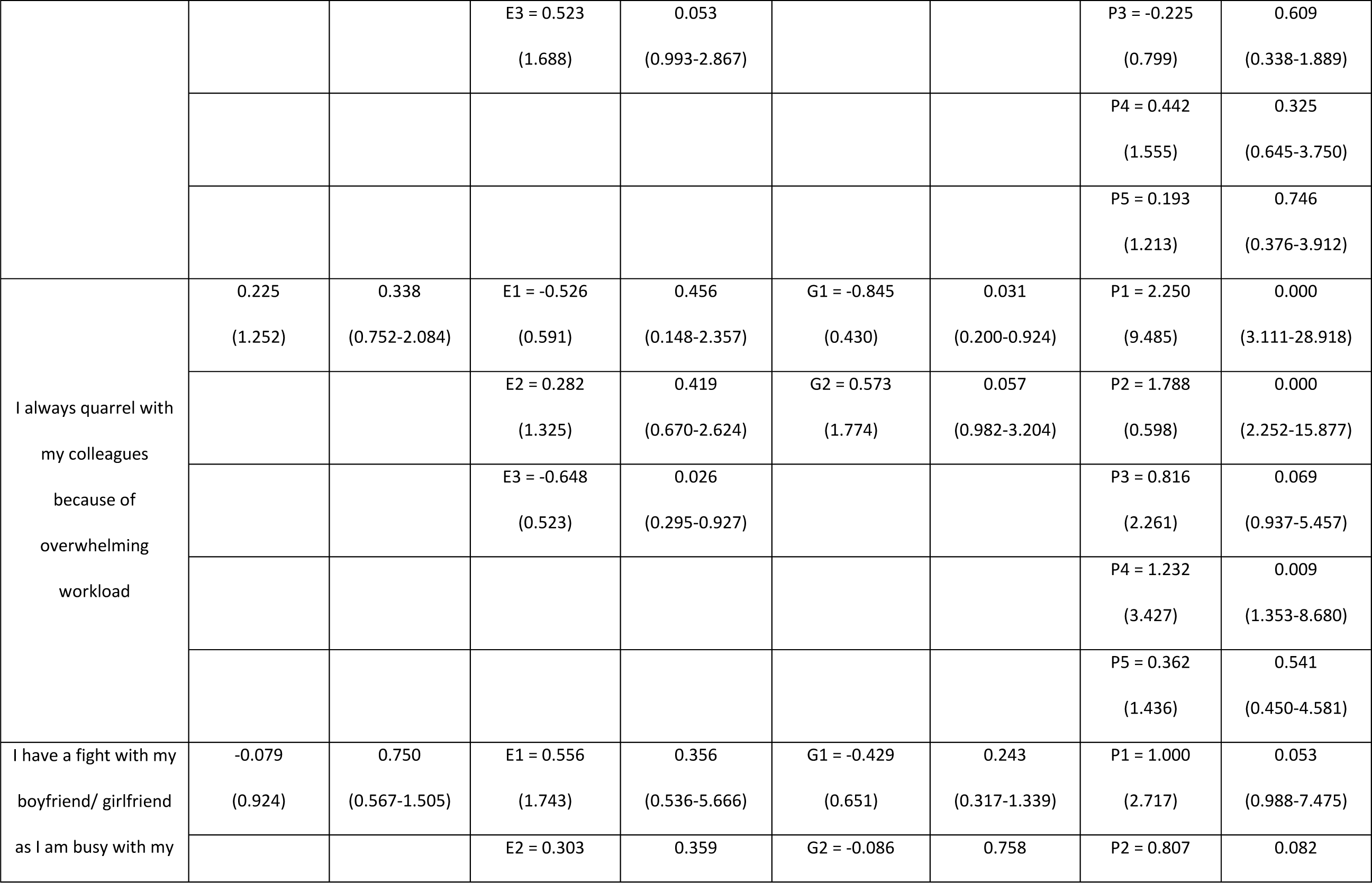
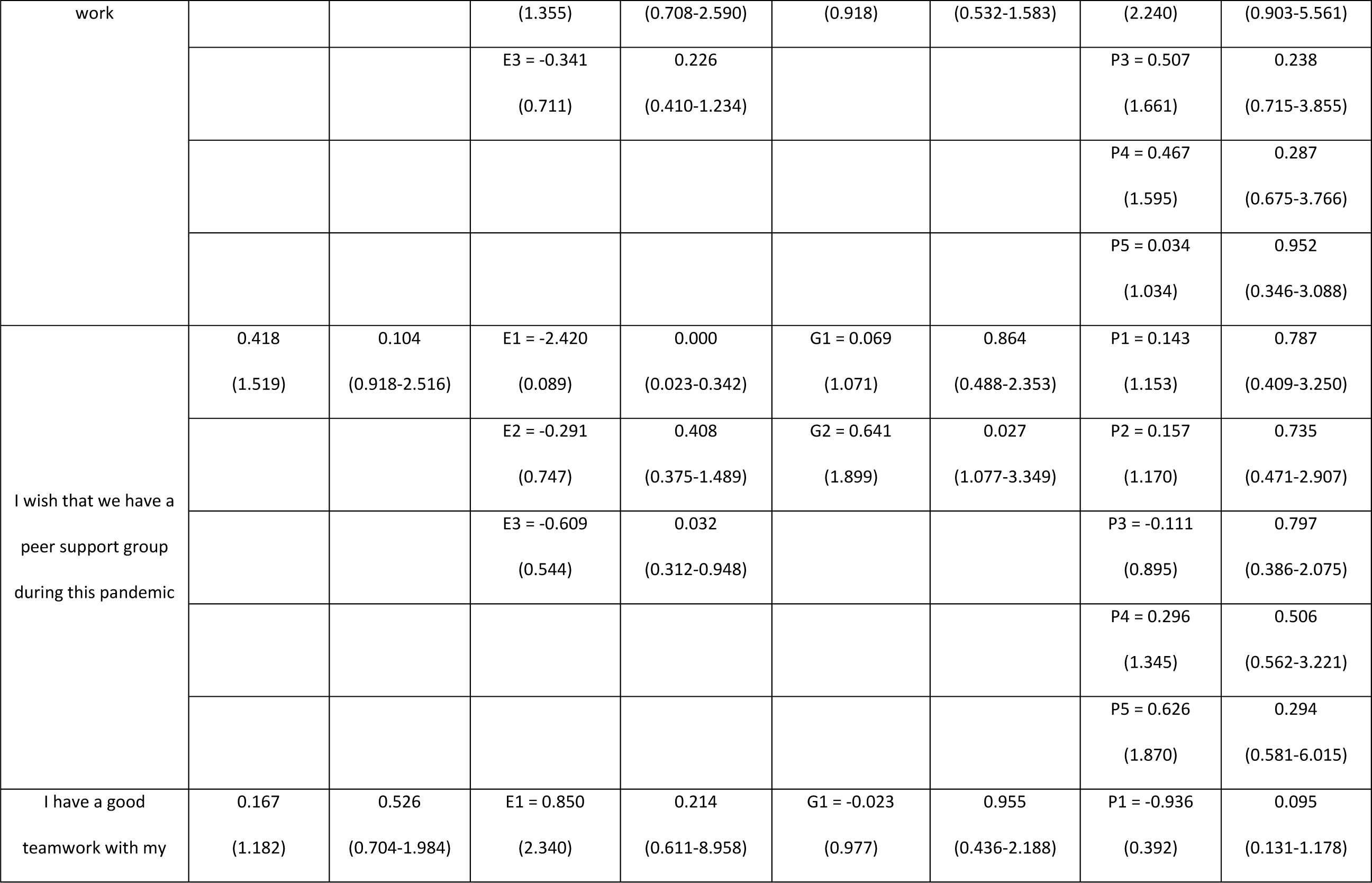
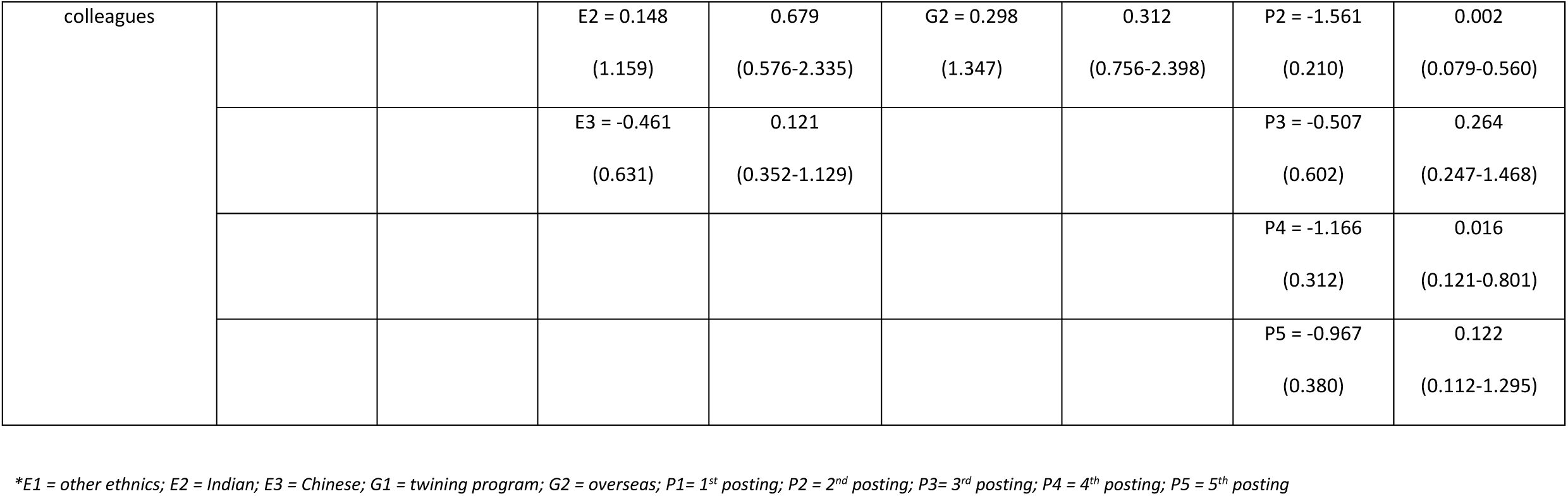
Association between Demographic Factors with Social/ Relationship Handling

### Physical Health

From the analysis, majority of the medical interns feel that they have not enough rest and exercises: males do not think they have enough exercise [p = 0.037, OR = 0.599, (95% CI 0.370 – 0.970)] if compared to girls; while Chinese medical interns think they do not have enough rest [p = 0.048, OR = 0.570 (95% CI 0.326 - 0.996)] Contrary, the medical interns who are graduated from twinning program think that they have enough rest and exercise. [p = 0.01, OR = 2.868 (95% CI 1.287 – 6.392) and p = 0.017, OR = 2.592 (95% CI 1.187 - 5.662) respectively] Viewing from the number of posting, first year junior medical interns do not think they have enough rest [ (P1) p = 0.014, OR = 0.278 (95% CI 0.100 - 0.774) / (P2) p = 0.032, OR = 0.352 (95% CI 0.136 - 0.912)] and they experience hunger most of the time [ (P1) p = 0.003, OR = 4.658 (95% CI1.686 - 12.872)/ (P2) p = 0.004, OR = 3.995 (95% CI 1.553 - 10.279)/ (P3) p = 0.012, OR = 3.015 (95% CI 1.278 −7.115)] While third and fourth posting interns think they have enough exercise. [(P3) p = 0.004, OR = 3.307 (95% CI 1.459-7.494) / (P4) p = 0.009, OR = 3.117, (95% CI 1.333-7.292)]. On the other hands, they feel they are burden with heavy workload, especially the first posting [ (P1) p = 0.023, OR = 3.377 (95% CI 1.182 - 9.648)/ (P4) p = 0.047, OR = 2.42 (95% CI 1.013 - 5.781)].

### Emotion/ Psychological Coping

For the statement “If given a chance to turn back time, I would not choose to be a doctor”, majority of male do not regret becoming a doctor [p= 0.032, OR = 0.586 (95% CI 0.359-0.956)]. During this pandemic crisis, Chinese medical interns are scared to go to work [p = 0.030, OR = 0.546 (95% CI 0.316 - 0.944)] while Indian and other ethnics groups are worried to be infected with COVID during their works [(Indian) p = 0.017, OR = 0.217 (95% CI 0.062-0.763)/ (Others) p = 0.000, OR = 0.290 (95% CI 0.151 - 0.555)] Junior medical interns are more worried and depressed than others while working during this pandemic situation: medical interns who are from second posting feel depressed [p = 0.043, OR = 2.619 (95% CI 1.029-6.668)] while third posting worried they might get infected with the virus [p =0.033, OR = 0.403 (95% CI 0.175 - 0.929)]. There is no association between their psychological coping and where they attained their medical degree.

### Social/ Relationship Handling

During this pandemic period, even though there is a change in our healthcare system, most of the Chinese medical interns do not quarrel with their colleagues during work [p = 0.026, OR = 0.523 (95% CI 0.295 - 0.927)] and same goes to those who graduated from twinning program [p= 0.031, OR = 0.430 (95% CI 0.200 - 0.924)] However, based on the number of postings, most of them quarrel with their colleagues due to overwhelming workload [(P1) p = 0.000, OR = 9.485 (95% CI 3.111 - 28.918)/ (P2) p = 0.000, OR = 0.598 (95% CI 2.252 - 15.877)/ (P4) p = 0.009, OR = 3.427 (95% CI 1.353 - 8.680)]. At the same time, Chinese and other ethnics do not think there is a need of peer support group [(Chinese) p = 0.032, OR = 0.544 (95% CI 0.312 - 0.948) / (Others) p = 0.000, OR = 0.089, 95% CI 0.023 - 0.342)] but those who are graduated from overseas think a peer support group will be beneficial [p = 0.027, OR = 1.899 (95% CI 1.077-3.349)]. Medical interns in first posting think they have lesser contact with their friends during this period [p = 0.022, OR = 3.355 (95% CI 1.191 - 9.447)]

## Discussion

In general, HO are medical interns who require at least two years of medical training before they are allowed to practice medicine in Malaysia.[6] Being part of the training is stressful as they must go through different fields and learn to manage patients in respective fields within limited time. Furthermore, they have pressures from superiors, expectation from patients and requirements to be fulfilled by performing numbers of basic procedures within that short period of time. [6, 12-14]

COVID-19 pandemic has affected all HCW greatly in terms of routine ward rounds, elective operations and clinic arrangement, thus disrupts our current healthcare system. [15, 16] Additionally, they are suffered from immense stresses as they are worried of contacting the virus while treating patients. [11, 17] In addition, educational training and medical teaching session have been ceased until further notice. They have inadequate cases and procedures to observe; facing lack of experience and hands-on clinical skills; being re-allocated depending on capacity and workload of the hospital.[8, 9] Due to the tremendous uncertainty during this pandemic crisis, medical interns as the junior most doctors face far more stresses than other HCW.

### Physical Health

Many medical interns feel like they do not have enough rest, food and exercise. This result shows that they are busy and occupied with work until they could not have adequate rest or even meals. On the other hands, the reason they lack exercises could be due to busy work life during the day which exhaust them physically or due to COVID pandemic where recreational activities and gyms are forbidden. In addition, they are burdened with heavy workload while working during this pandemic period. Among medical interns, they are divided into small groups while attending patients to minimise close contact. This strategy aims to prevent cross transmission between HCW but unfortunately reduces the numbers of attending doctors per shift thus lead to relatively high workload. Expectedly, first posting of medical interns are affected the most as they are new to their jobs. As a fresh graduate who just entered a new working environment, not only they need to manage patients and perform ward works but they require extra effort to equipped themselves to deal with pandemic. This pandemic crisis causes a surge of patients’ loads which might exceed their capabilities thus encumber them further.

However, medical student who are graduated from twining program (both local and overseas universities and colleges) beg for differ where they feel that they have enough rest and exercises during this period. By attending universities from two different countries and two different cultures might allow the medical interns to adapt to new environment more easily and readily if compared with others.

### Emotion/ Psychological Coping

Majority of the medical interns are worried of contacting COVID-19 while working in a COVID centre. Some of them are in fear and even depressed to head to work during this pandemic situation. The results are expected as many literatures have showed that the psychological impacts of pandemic towards HCW are significant.[2, 8, 10, 17-21] During this pandemic season, HCW experiences a complex emotion such as fear, worry, anxiety and even depression. To work under such circumstances, mental health of HCW will be affected inevitably. For a HCW to fight against a virus with no cure at the moment and to deal with pressures from patients and communities, unimaginable stress could be exerted to them. Besides, fear of local transmission within HCW and worry of transmitting the virus to their families heighten their tensions inevitably. This could be the aftermath effect of pandemic crisis which can eventually lead to acute psychological distress, burnout, and posttraumatic stress.[22]

Interestingly, most males do not regret of being a doctor. [23, 24] This could be due to our cultural influence that male should always be firm and decisive to carry out their responsibilities with their strong will.

### Social/ Relationship Handling

Since medical interns spend most of their time working, relationships with colleagues are unavoidably affected during this period. Most of the junior medical interns’ have conflicts with their colleagues during this period due to overwhelming workload. This manifestation could be one of the signs that medical interns are under work stress. [10] They could be overworked and fatigue which lead to miscommunication and misunderstanding. Without trust, spirit of teamwork and harmony could not be established and lead to unhappiness and dissatisfaction in workplace.

On the other hand, some feel like that they have lesser contact with their friends during this period even with the use of telecommunication devices. Possibility that they are occupied with their heavy workload and do not have time to socialise. This will indirectly cultivate antisocial and indifference behaviours among medical interns.

As for peer support group, it seems to be beneficial for most of the interns who graduated from overseas but Chinese and other ethnics choose to differ. For international medical students to adapt and survive overseas, peer support and social support are commonly introduced. Peer support plays an important role in holding them together to overcome this difficult time and lessen their tensions by sharing their thoughts with others.

### Limitation

1. This is a pilot study where the questionnaire is not validated. The questionnaire is created based on problems encountered by medical interns during their daily routines.
2. As for the sample size, only 236 medical interns participate in this study with the initial calculated sample size of 378. This contributes to 63% of response rate. [25, 26] There are few problems identified which are some of the medical interns have finished their internship and had switched to other hospitals or awaiting placement; some of them are in the transition of switching departments; invalid email addresses; possibility of busy working schedules or being too preoccupied with their routine works.
3. In addition, this is a self-administered questionnaire in anonymity where repetition of entry is possible. Follow-up study could be helpful in assessing the progression of medical interns after COVID-19 pandemic subsides.

## Conclusion

COVID-19 pandemic has invaded many countries and paralysed most of the healthcare systems. During this challenging period, medical interns who are the junior most doctors suffered significantly especially their physical health, emotion states and social lives.

To ensure physical needs of medical interns are fulfilled, a sufficient protected time should be given to them, such as time for break and meals in between their works without neglecting their duties. During pandemic period where movement restriction order is enforced, restaurants and food suppliers are forbidden to open at night; to provide meals during their oncall or night shifts might solve their problems. Without proper rest and energy, not only their health but patients’ safety will be jeopardised.

Psychological impact of the pandemic on HCW is well reported in many literatures, similar results show in medical interns too. The cause of it mainly due to insecurity which most of the HCW feel when facing pandemic situation: lack of latest medical information regarding COVID-19 and insufficient medical resources and personal protective equipment (PPE) while treating patients. To complicate the situation, lack of psychological support amongst medical interns often drift them away when they are at the edge of breakdown. [10, 20] Early detection of mental instability warrants an early referral to psychiatrists or counsellors. Implementation of counselling services and peer support groups should be established to overcome mental health issues which are encountered by medical interns.[27, 28] With the aid of these measures, working relationships with colleagues shall be improved and unnecessary conflicts could be avoided.

In a nutshell, the impact of COVID-19 amongst medical interns is significant and should be addressed appropriately. Their overall well-being should be protected and preserved without jeopardise their training and preparation for pandemic crisis in the future.

## Data Availability

All data are available in the manuscript.

## Abbreviations

COVID-19: Coronavirus Disease 2019
HCW: healthcare workers
UMMC: University Malaya Medical Centre
OR: Odd Ratio

## Acknowledgement

I sincerely would like to thank everyone who has contributed to this study. A heartful gratitude to all parties in helping to make this study a success:

1. House Officer Coordinators from different departments in UMMC
2. Human Resource Officers UMMC
3. Dr Retnagowri A/P Rajadram, Senior Lecturer, Research Unit, Department of Surgery, UMMC, Malaysia
4. Professor Dr April Roslani, Head of Department of Surgery, UMMC, Malaysia
5. Professor Dr. Tunku Kamarul Zaman Bin Tunku Zainol Abidin, Director of UMMC, Malaysia

